# *Triatoma dimidiata,* domestic animals and acute Chagas disease: A 10 year follow-up after an eco-bio-social intervention

**DOI:** 10.1101/2025.03.11.25323671

**Authors:** Jose G. Juarez, Andrea M. Moller-Vasquez, Maria Granados-Presa, Pamela Pennington, Norma Padilla, Sujata Balasubramanian, Lisa D. Auckland, Elsa Berganza, Luis Alvarado, Henry Esquivel, Ranferi Trampe, Louisa Messenger, Celia Córdon-Rosales, Gabriel L. Hamer, Sarah A. Hamer

**Affiliations:** Sustaianble Sciences Institute, Oakland, California; Centro de Estudios en Salud, Universidad del Valle de Guatemala, Guatemala; College of Veterinary Medicine & Biomedical Sciences, Texas A&M University, College Station, US; Departamento de Epidemiología de la Dirección Departamental de Redes de Servicios Integrados de Salud Jutiapa, Ministerio de Salud Pública y Asistencia Social, Guatemala; Sección de Vectores de Jutiapa de Redes de Servicios Integrados de Salud Jutiapa, Ministerio de Salud Pública y Asistencia Social, Guatemala; Parasitology and Vector Biology Laboratory (PARAVEC Lab), School of Public Health, University of Nevada, Las Vegas, NV, USA; Department of Environmental and Occupational Health University of Nevada Las Vegas, Las Vegas, NV, USA; Department of Entomology, Texas A&M University, College Station, US

**Keywords:** Triatomines, Follow-up, Chagas disease, treatment, persistent, acute cases

## Abstract

**Introduction:** *Trypanosoma cruzi*, the causative agent of Chagas disease, is primarily transmitted by triatomine insects, including *Triatoma dimidiata*. In Central America, vector control programs have significantly reduced transmission; however, certain regions, such as Comapa, Jutiapa, Guatemala, continue to experience persistent *T. dimidiata* infestation. This study presents a 10-year follow-up assessment of triatomine infestation, *T. cruzi* infection, and acute Chagas disease cases after an eco-bio-social intervention.

**Methods:** Between June and August 2022, entomological surveys were conducted in four communities of Comapa. Seventy six households were systematically searched for triatomines using the one-person hour method, which were collected and processed for *T. cruzi* detection using qPCR. Bloodmeal analysis was performed to assess host feeding patterns. Dog samples and environmental DNA from household surfaces were also processed for *T. cruzi* detection. Additionally, surveillance for acute Chagas disease cases was carried out in collaboration with the Ministry of Health.

**Results:** Persistent infestation of *T. dimidiata* was observed across all communities, with infestation rates ranging from 17–38% and colonization levels between 9–29%. The mean household triatomine density remained low, suggesting a possible reduction in transmission risk. A total of 86 triatomines were collected, of which 26% tested positive for *T. cruzi* (all TcI strain). Amplicon deep sequencing analysis from triatomines identified seven vertebrate species and one insect family as hosts upon which triatomines have previously fed, with chickens being the most common blood source (occurring in 57% of triatomines), along with rats, dogs, humans, cats, pigs, ducks, and one genus of cockroach. Of the 132 dogs processed 22% were positive for *T. cruzi* (all TcI). One acute Chagas disease case detected in a child in 2015 remained seropositive in 2022, emphasizing the need for continued surveillance.

**Conclusions:** Despite multiple interventions over a decade, *T. dimidiata* infestation remains high in Comapa with sustained evidence of actue disease in humans, necessitating continued vector control efforts. The persistence of *T. cruzi* transmission among triatomines and dogs and the predominant role of chickens in supporting the vector population highlights the need for innovative control strategies including those that target domestic animals to mitigate Chagas disease risk.

## Introduction

Chagas disease, caused by the parasite *Trypanosoma cruzi*, is a neglected tropical disease that infects around 7 million people worldwide, with most cases found in Latin America [1]. The disease is primarily transmitted by insects of the family Reduviidae[1]. Despite being considered one of the most neglected tropical diseases and having one of the highest disability-adjusted life years (DALYs) of any infectious disease in the Americas [2,3], its control still heavily relies on traditional vector control strategies focused on indoor residual spraying due to the lack of effective vaccines and drug therapies with lower toxicity and higher efficiency [4].

The *T. cruzi* infection has a 7-15-day incubation period after initial infection, followed by an acute phase that lasts an average of eight weeks, which can be diagnosed through blood Strout or blood smear examination to detect circulating parasites. However, only 1-2% of cases report symptoms at this stage, including chagomas, (inflammatory reactions at the inoculation site) and the Romaña sign (swelling of the eyelid), making acute case diagnosis extremely rare [5,6]. Chronic disease manifestations may occur, including Chagas cardiomyopathy, occur among a subset of infected individuals [7]. In Central America, vector control interventions based on Insecticide Residual Spraying (IRS) carried out during the first decade in 2000 are responsible for reducing vectorial transmission by 94% [8]. However, there are still regions where persistent indoor infestation remains high with *Triatoma dimidiata*.

Comapa, Jutiapa in Guatemala is a region of sustained transmission, with indoor infestation of *T. dimidiata* ≥15%, exeding the 8% threshold consider necessary for sustainable disease control, even after multiple rounds of indoor residual spraying [9]. In 2011, an entomological baseline survey of *T. dimidiata* confirmed high infestation indices across multiple communities of Comapa [9]. In 2012, a multistakeholder community-based intervention focused on ecological, biological and sociological risk factors [9] was developed and successfully adopted by households for the management of rodents in the peridomicile to control Chagas disease [10,11]. In addition, an intervention focused on house improvement was developed in the same area [12]. Results from a 2015 seroprevalence study showed that seropositivity in school-age children had decreased from 9.2% before the mass insecticide applications to 1.6% over a decade after the mass insecticide applications were completed [13]. However, beyond understanding how an intervention might be adopted and implemented by community members [14], there is a need to evaluate the risk inhabitants might face in these communities with persistent *T. dimidiata* infestation and how other entomological indices, such as vector household density, could provide a more nuanced evaluation of household risk. In this study, we report the triatomine infestation, *T. cruzi* infection and bloodmeal analysis results for four communities in Comapa, Jutiapa, as well as the level of infection in domestic dogs and occurrence of human disease.

## Methods

### Ethic statement

This research was reviewed by the Research Ethics Committee of Centro de Estudios en Salud at Universidad del Valle de Guatemala (UVG) which classified it as “Research not involving human subjects” (Protocol No. 270-05-2022). It was also reviewed by the Institutional Animal Care and Use committee of UVG (CEUCA-UVG) and was approved under protocol number I-2022(3)A. Additionally, it was approved by the Texas A&M Universitýs Institutional Animal Care and Use Committee (IACUC 2022-0001 CA). Human acute Chagas disease cases were part of the national surveillance program of the Guatemalan Ministry of Health. All information shared with UVG had no personal identifiers that could be link to patients. Verbal consent was obtained from homeowners for entomological surveillance. Signed conset was obtained from the parent of the acute Chagas disease case, for eye image publication.

### Study sites and sample size

The information regarding the study communities and prior intervention has been previously published [10]. Briefly, this research was conducted in the department of Jutiapa, municipality of Comapa, located in southeastern Guatemala (Figure 1). The research was conducted in four communities of Comapa from June to August 2022. We randomly selected four communities; two communities were previously control (Buena Vista=BV and El Comalito = EC) and two communities were previously part of an intervention receiving education regarding risk factors and environmental management (San Antonio=SA and Santa Barbara=SB) [10]. Our previous study documented >15% of the households with *T. dimidiata* infestation, out of a total of 18 communities that took part in an eco-bio-social intervention for the control of Chagas disease [10,11,16]. As previously done, we intended to survey the same 24 households (as per the guidelines of the Guatemalan Ministry of Health [17]) from the original selection of 2012 [10], to provide us with information regarding *T. dimidiata* infestation and *T. cruzi* infection rates 10 years after the intervention took place. Additionally, a community member from El Anonito requested entomological surveillance help in her household where many triatomines occurred, results for this household are presented separately in Supplementary Results.

**Figure 1.**
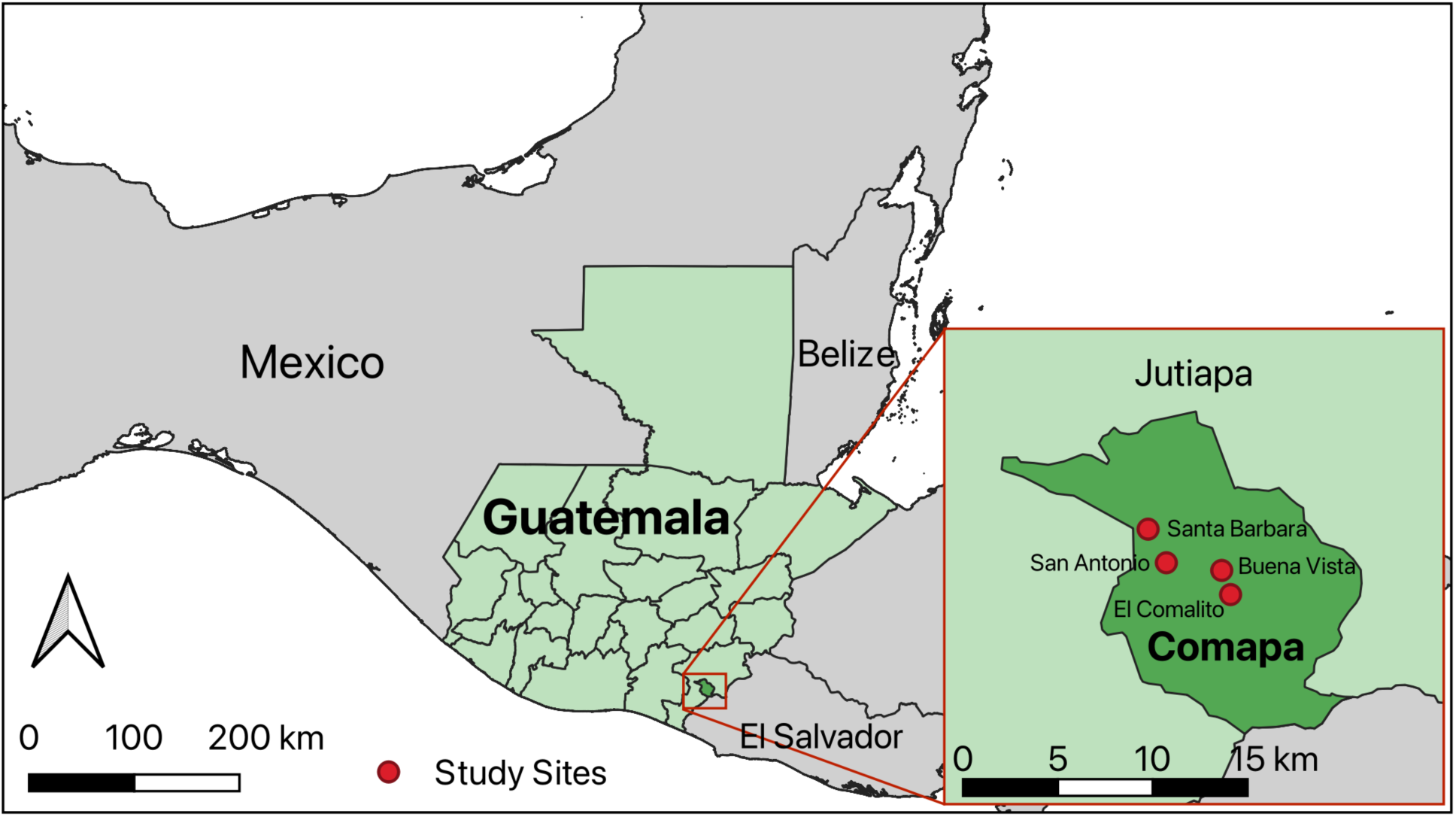
Triatomine collections sites in Comapa, Jutiapa, Guatemala (2022). Map was generated using Quantum GIS (QGIS 3.30) using freely available administrative boundaries.

### Entomological surveillance

The search for triatomines was conducted by our research team alongside Ministry of Health (MoH) personnel following the one person-hour methodology [17]. The highly trained MoH personnel thoroughly searched all premises of the household, outside walls and peridomicile structures for triatomines. Search time could extend depending on the number of rooms, items required to be moved and/or triatomines found. Triatomines were classified by developmental stage, feeding status, sex, and species. All specimens were stored in 70% ethanol, samples were later moved into RNAlater® (Invitrogen) and stored at -80°C until further processing for *T. cruzi* infection and bloodmeal analysis. Triatomines collected in the additional house from El Anonito were kept alive and taken to the laboratory at UVG. Adult triatomines from El Anonito were not processed since they were used to establish a *T. dimidiata* colony; all instars were processed. We present three entomological indicators i) infestation (% of households with triatomines), ii) colonization (% of households with triatomine instars) and iii) vector density (average of triatomines found in a household).

### Environmental DNA

Environmental sampling to detect eDNA of *T. cruzi* [18] was conducted using two polyester swabs, which were wiped over an approximate 60 cm × 60 cm area of the wall and floor adjacent to the bed in each household. Each surface was swabbed for approximately 15 seconds, and swabs were subsequently placed into a collection tube containing ∼250uL of RNAlater® Samples were then frozen until further processing. DNA extraction was performed using the Omega E.Z.N.A. Tissue DNA Kit, following the manufacturer’s protocol with a modification to the elution step. Instead of a single elution step, a two-step elution was used: 25 µL of elution buffer was added, followed by centrifugation, then an additional 25 µL was added before a second centrifugation, resulting in a final elution volume of 50 µL. Quantitative PCR (qPCR) was to detect the presence of *T. cruzi* DNA was performed as described below.

### Triatomine processing

All specimens were handled under Biosafety level 2 hoods, with pictures taken and length measurement recorded. We dissected the body and separated the hindgut and remaining body into separate tubes for instars 4 and 5, and adults. This procedure involved dipping the specimen in sterilized water for 10 sec and then 15 sec in 50% bleach, followed by cutting the connexivum to remove the guts. Records were kept for feeding score of each specimen as follows: 1: No blood, no guts; 2: Guts found, no feeding; 3: Small traces of feeding; 4: Evidence of feeding, but limited; and 5: Evidence of feeding, engorged. Instars 3 and below were scored using a less revolved scale of fed, slightly fed or unfed.

We used the MagMax-96 DNA Multi-Sample Extraction kit (Applied Biosystems) for extraction of total nucleic acid from the hindgut of individual triatomines. If tissue sample was above 10mg, the volume of proteinase K would be doubled. To lyse the samples, they were left overnight at 55°C in a shaker. Samples were further processed following the procedure previously described [19].

### Dog sampling and processing

Dogs owned by household residents were restrained and a muzzle was placed. The venipuncture site was disinfected with 70% ethanol and up to 5 mL of blood was collected into EDTA tubes and initially stored at 4°C followed by centrifugation and -80°C storage of the serum and blood clot. All personnel involved in dog handling and sample collection were vaccinated against rabies. For each dog data for sex, age and breed were recorded. Ectoparasites from dogs has already been presented and analyzed in a separate manuscript [20].

### *Trypanosoma cruzi* detection and strain genetic characterization

We performed quantitative real-time PCR (qPCR) to detect the presence of *T. cruzi* in all samples (DNA from house swabs, triatomines and dog blood) using a Stratagene MxPro3000 (Agilent Technologies). All samples were processed using 3μl of previously extracted DNA, primers at a volume of 0.3μl (Cruzi 1: 5′-ASTCGGCTGATCGTTTTCGA-3; Cruzi 2: 5′-AATTCCTCCAAGCAGCGGATA-3; and Cruzi 3: 5′-Fam-CACACACTGGACACCAA-NFQ-MGB-3) [21], probe concentration of 0.45μM, and a mix of PCR FailSafe enzymes with PreMix E. The amplification target was a 166bp region of repetitive microsatellite nuclear DNA detected using TaqMan probes. Negative controls were DNA free water and positive controls were DNA recovered from *T. cruzi* positive *Macaca fascicularis*. Dog blood samples were additionally processed for antibody detection of *T. cruzi* with an off-label use of the rapid serological test, Chagas Stat-Pak Assay (Chembio Diagnostic Systems, US) following manufacturer proceedures[22].

All samples that tested qPCR-positive for *T. cruzi* were subjected to genetic characterization of the parasite using a RT-qPCR of the SL-IR gene of *T. cruzi* with differential probes for each DTU[23,24]. The protocol used a multiplex PCR kit (Qiagen, Germany), with the oligonucleotides, sequence, and concentration in Supplementary Results Table 1. Samples were processed at a final volume of 18μl by reaction with positive controls for the DTUs[24]. Samples without a DTU identification were rerun at a 1:10 concentration and if no positive result was obtained, they were left as untypable.

**Table 1.** Household triatomine indices for *T. dimidiata* collections and *T. cruzi* infection in four communities of Comapa, Jutiapa.

| Variable | Overall | Community |  |  |  |
| --- | --- | --- | --- | --- | --- |
|  |  | BV<sup>a</sup> | EC<sup>a</sup> | SA<sup>b</sup> | SB<sup>b</sup> |
| **Households** | **N= 76** | **N= 24** | **N= 5** | **N= 23** | **N= 24** |
| **Infestation** |  |  |  |  |  |
| Overall | 20 (26%) | 9 (38%) | 1 (20%) | 4 (17%) | 6 (25%) |
| Domicile | 14 | 7 | 0 | 4 | 3 |
| Peridomicile | 7 | 2 | 1 | 1 | 3 |
| **Colonization** |  |  |  |  |  |
| Overall | 14 (18%) | 7 (29%) | 1 (20%) | 2 (9%) | 4 (17%) |
| Domicile | 8 | 5 | 0 | 2 | 1 |
| Peridomicile | 6 | 2 | 1 | 0 | 3 |
| **Life Stage** | **N= 86** | **N= 53** | **N= 3** | **N= 21** | **N= 9** |
| 1<sup>st</sup> instar | 9 (10%) | 6 (11%) | 0 (0%) | 3 (14%) | 0 (0%) |
| 2<sup>nd</sup> instar | 25 (29%) | 17 (32%) | 2 (67%) | 4 (19%) | 2 (22%) |
| 3<sup>rd</sup> instar | 16 (19%) | 7 (13%) | 0 (0%) | 7 (33%) | 2 (22%) |
| 4<sup>th</sup> instar | 1 (1.2%) | 1 (1.9%) | 0 (0%) | 0 (0%) | 0 (0%) |
| 5<sup>th</sup> instar | 14 (16%) | 9 (17%) | 1 (33%) | 1 (4.8%) | 3 (33%) |
| Female | 9 (10%) | 6 (11%) | 0 (0%) | 3 (14%) | 0 (0%) |
| Male | 12 (14%) | 7 (13%) | 0 (0%) | 3 (14%) | 2 (22%) |
| **Location** |  |  |  |  |  |
| Domicile | 62 (72%) | 37 (70%) | 0 (0%) | 20 (95%) | 5 (56%) |
| Peridomicile | 24 (28%) | 16 (30%) | 3 (100%) | 1 (4.8%) | 4 (44%) |
| ***T. cruzi*** |  |  |  |  |  |
| Negative | 64 (74%) | 39 (74%) | 0 (0%) | 19 (90%) | 6 (67%) |
| Positive | 22 (26%) | 14 (26%) | 3 (100%) | 2 (9.5%) | 3 (33%) |
| **Strain** |  |  |  |  |  |
| Tc1 | 20 (91%) | 12 (86%) | 3 (100%) | 2 (100%) | 3 (100%) |
| Untypable | 2 (9.1%) | 2 (14%) | 0 (0%) | 0 (0%) | 0 (0%) |

<sup>a</sup>= historic control community; <sup>b</sup>= historic intervention community

### Triatomine bloodmeal analysis

Bloodmeal analysis was based on deep sequencing of amplicons generated for a region of the vertebrate 12S rRNA mitochondrial locus amplified from the extracted DNA of triatomines [25–27]. Amplification of the 145 bp fragment was done in duplicate for every sample. Forward and reverse primers included dual identical barcodes [28]. Samples were handled in clean and disinfected biosafety cabinets and in separate pre- and post-PCR areas of the laboratory to minimize contamination. Samples with appropriate sized bands visible on gel electrophoresis were submitted to the Texas Institute for Genome Sciences (https://genomics.tamu.edu) for library preparation using xGen^TM^ ssDNA & Low Input DNA Library Prep kit (Integrated DNA technologies, Coralville, IA, USA) and sequencing on the Illumina NextSeq 2000 (Illumina, San Diego, CA, USA). Barcodes were trimmed and demultiplexed using Cutadapt 5.0 [29]. Sequences were imported into Qiime2 [30] and primers were trimmed, sequences were merged and denoised at minimum length of 100 bp. Representative sequences were matched to taxa using NCBI BLAST (National Center for Biotechnology Information, US National Library of Medicine) with MegaBLAST with GenBank database (https://www.ncbi.nlm.nih.gov/genbank/)[31]. Only host species appearing in both replicates and showing more than 500 reads were retained. Species level identification of hosts was accepted at 99-100% identity to the database match. Lower percent identity scores (below 99%) were categorized to genus level. If multiple matches were found with the same percent identity, the host was identified up to the lowest common taxon.

### Acute Chagas disease surveillance

Since the time of our initial intervention project [9], the MoH stablished an active acute Chagas disease surveillance project between March 2013 and June 2015. All Chagas disease suspected acute case (any person with Romaña sign or with chagoma) detected by community members, MoH personnel or UVG were referred to the epidemiology unit of Jutiapa. Households with a suspected acute case were visited by vector control and laboratory personnel from the MoH. The household was searched for triatomines using the hour/person method and a 5 ml peripheral blood sample was collected from all children in the household for parasitological diagnosis using the Strout (< 7 years of age) technique [32]. Collected triatomines were transported alive to UVG laboratories for microscopic confirmation of *T. cruzi*. Triatomines that arrived alive had their hindgut punctured and 100μl of phosphate-buffered saline solution was used to wash the punctured area to extract a diluted portion of the fecal matter. Microscopic confirmation of *T. cruzi* was done using a 600X magnification by phase contrast microscopy. Positive children were tracked to follow-up on their Chagasic status during November 2022.

### Statistical analysis

Initial descriptive analyses were performed for data exploration and normality was evaluated using Shapiro-Wilk normality test. Overall entomological comparisons between domicile and peridomicile household infestation and colonization were analyzed using a Pearson’s Chi-square tests with Yate’s continuity correction at a p-value of <0.05. Triatomine abundance between domicile and peridomicile by community were analyzed using a Wilcoxon Rank Sum test. All analysis and graphs were generated with R 4.4.1 [33]. Due to small sample size, we avoided comparing El Anonito in the analysis.

## Results

### High infestation and colonization levels of *Triatoma dimidiata*

In the four communities surveyed, the number of homes with follow-up were 24 houses in both Buena Vista and San Antonio, 23 houses in Santa Barbara and five houses in El Comalito, for a total of 76 households (Table 1). We observed a high infestation level with *T. dimidiata* that ranged from 20-38% in the control (BV and EC) and 17-25% in intervention communities (SA and SB). Additionally colonization of households was between 20-29% in control and 9-14% in intervention communities. We did not observe any statistical difference between domicile and peridomicile household infestation and colonization. We collected a total of 86 *T. dimidiata* specimens (Instars= 65; Adults=21) across all four communities. BV (control) had the highest abundance of total triatomines found with 53 specimens (Instars=40; Adults=13) and a mean density per household of 2.21 (SD=3.99) triatomines. SA (intervention) had a total of 21 triatomines (Instar=15; Adult=6) and a mean density of 0.91 (SD=3.54). SB (intervention) had a total of 9 triatomines (Instar=7; Adult=2) and a mean density of 0.37 (SD=0.77). Lastly, EC (control) had a total of 3 triatomines (Instar=3; Adult=0) and a mean density of 0.6 (SD=1.34). We observed wide distribution of instars and adults in both domicile and peridomicile environments (Figure 2A). Male triatomines collected in the domicile were 56% compared to females which was 45%, instars collected in the domicile varied from 40 to 60% of overall collections, with no statistical difference observed.

**Figure 2.**
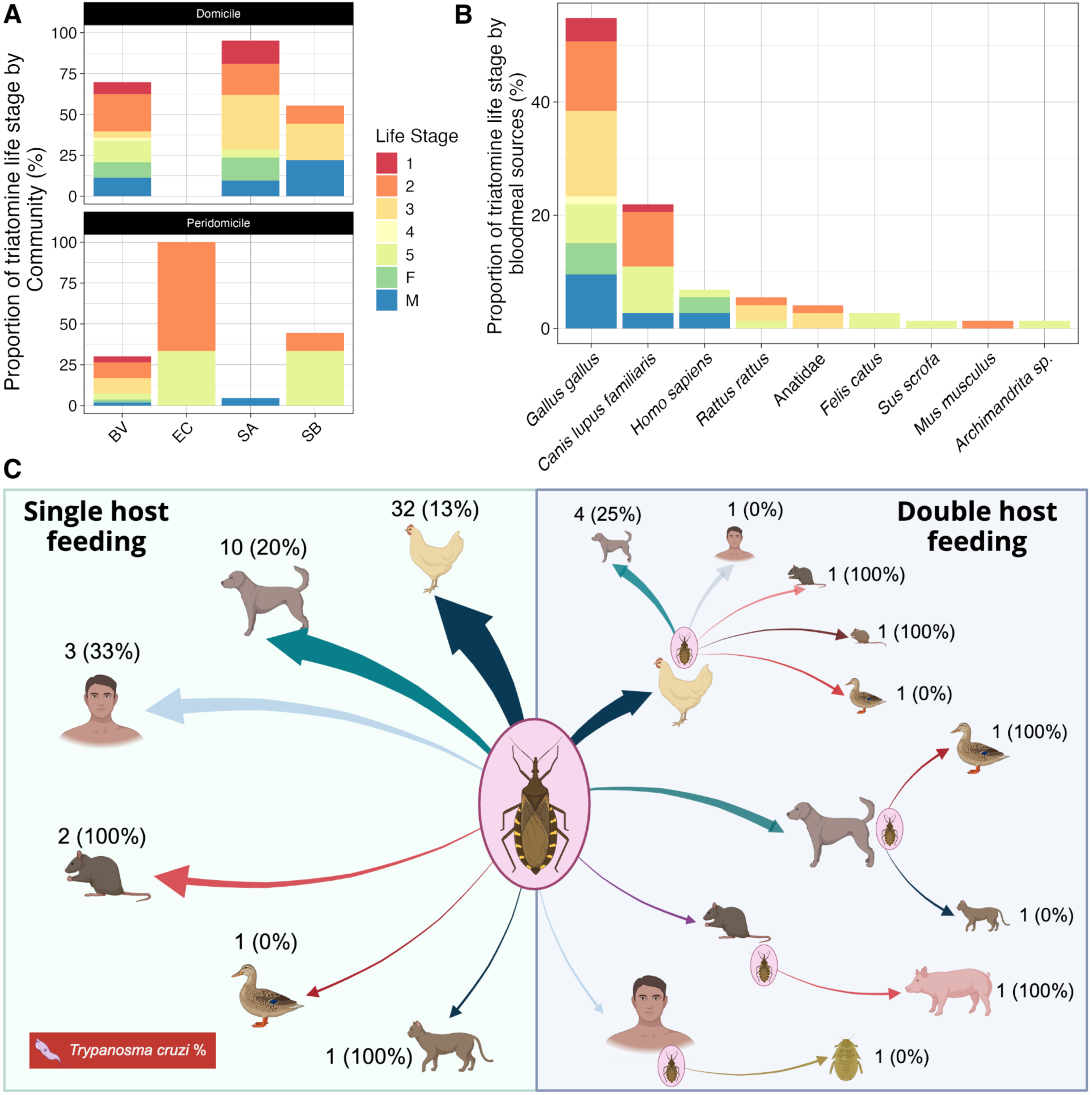
*Triatoma dimidiata* bloodmeal analysis in Comapa, Jutiapa, Guatemala (2022). A) Relative abundance of triatomine domicile and peridomicile collections by life stages and communities. B) Relative abundance of host feeding sources of triatomines by life stages. C) Bloodmeal source for single and double host feeding, with percentage of positive *T. cruzi* bloodmeals. The size of arrow is proportionate to number of observed bloodmeals from a given host. Figure 2C created with BioRender.

### *T. dimidiata* and dog infection with *T. cruzi* TcI strain

Of the 86 triatomines processed for *T. cruzi*, we detected *T. cruzi* DNA in 22 (26%), all of which came from 10 households. In BV (control) we detected six households with *T. cruzi* infected triatomines, two households in SA (intervention), and one household for both EC (control) and SB (intervention). We observed positive immatures as early as 1^st^ instar (1 in BV), 2^nd^ (7), 3^rd^ (2), and 5^th^ (5), and five adults (3 females and 4 males). We did not observe any statistical difference in infection of triatomines between domicile and peridomicile specimens by life stage. Additionally, the only *T. cruzi* DTU detected in the 20 specimens that were positively typed was TcI [24].

We blood-sampled a total of 132 dogs, of which 19 (14%) had blood that was qPCR-positive for *T. cruzi* and 21 (16%) that were antibody positive. When combined we observed 28 (21%) positive dogs by either or both methods. Of those qPCR-positive, the DTU was determined for 11 samples and all were TcI. Dogs as early as 2-month-olds were found positive for *T. cruzi*.

### Household *T. cruzi* environmental DNA detection

We detected a total of 6 (n=145) *T. cruzi* positive swabs, from 5 different households, with 1 household having both wall and floor swabbing positive. We were able to determine DTU of 2 samples as TcI. The floors adjacent to the edge of the bed had the highest number of positive samples, with a total of 4, while only 2 positive samples were detected on the walls.

### Bloodmeal analysis of *T. dimidiata*

Out of 86 triatomines, 61 showed amplicons of the appropriate size after PCR for the vertebrate 12S rRNA gene. Seven vertebrate hosts were identified by species (*Canis lupus familiaris*, *Felis catus*, *Gallus gallus*, *Homo sapiens*, *Mus musculus*, *Rattus rattus*, *Sus scrofa*) and one by family (Anatidae) (Figure 2B). One sequence matched *Archimandrita* sp. (TD-3231-3), which is an invertebrate genus of cockroach; incidentally many Archimandrita species were observed in sampling locations at the time of collections (see Supplementary Results: SFigure1). Most triatomines (n=49) had sequences for a single host species and 12 showed two different hosts (Figure 2C). The most frequent meal was *Gallus gallus* (32 one-host bloodmeals and 8 two-host bloodmeals) followed by *Canis lupis familiaris* (10 one-host and 6 two-host bloodmeals) and *Rattus rattus* (2 one-host and 2 two-host bloodmeals) and. Human bloodmeals were identified from 3 triatomines as the sole bloodmeal host (one *T. cruzi* positive) and from 2 other triatomines along with *Gallus gallus* and *Archimandrita* sp.

### The tip of the iceberg: Acute Chagas disease

In March 2014, a 6 to 10 year old (y/o) boy from the community of San Juan, Comapa, was taken to the local health center since he presented Romaña’s sign and a week-long fever. The child was discharged, and a community health promoter recognized the symptoms and alerted vector control personnel of a probable acute Chagas disease case. A Strout test confirmed acute *T. cruzi* infection, and entomological inspection of the household revealed a single female *T. dimidiata* in the child’s bedroom, which tested positive for *T. cruzi* by rectal puncture and microscopy. In April 2015, a 1 to 5 y/o girl from Santa Bárbara, Comapa, exhibited Romaña’s sign and mild fever. A health educator trained in Chagas disease identification reported the case to Jutiapa’s epidemiology unit. A household inspection revealed 25 *T. dimidiata* specimens in the kitchen adjacent to the child’s bedroom. Thirteen specimens were transported alive to UVG, where three tested positive for *T. cruzi*. The child′s bloodsample also tested positive for *T. cruzi* via Strout and hemoculture. The remaining children in the household were also tested via Strout and ELISA with negative results. Both Chagas positive children were treated with nifurtimox under the supervision of a certified MoH nurse, who followed the protocol for Chagas disease management of the Guatemalan MoH [32]. In November 2022, the MoH conducted a follow-up to assess the children’s Chagasic status. Unfortunately, only the girl was located, and her ELISA test remained positive, which could be due to insufficient time for antibody waning, or possible reinfection, as *T. dimidiata* remains prevalent in the region.

## Discussion

The Chagas disease vector control program in Guatemala has been extremely successful with only a few pockets of persistent infestation remaining. One such pocket of persistent high infestation is in the southeastern region of Guatemala which has shown refractoriness to multiple vector control activities and interventions over the years. Our results showed that high infestation levels remain in the four communities evaluated, even more importantly, infection rates with *T. cruzi* were widespread in all the communities and in all life stages as early as 1^st^ instars. Dogs as young as 2-months-old were also found positive. *T. cruzi* DTU TcI remains the sole DTU detected in both triatomines and dogs, consistent with our prior study and suggesting no new DTU introductions [34]. Importantly, the follow-up of acute Chagasic children showcased that even after treatment, patient monitoring is critical to inform diagnosticians on how to proceed with the management of Chagas disease, and how the efforts of treatment might be thwarted by the persistence of triatomines in the region.

The 10-year follow-up evaluation of *T. dimidiata* in these four communities of Comapa, Jutiapa, showed that persistent infestation and colonization remain high in the region. Despite multiple efforts to reduce infestation levels through both traditional and ecological interventions[11], which have been widely accepted by community members [14], infestation persists. In contrast, a 20-year post-evaluation of an eco-health intervention in the community of La Brea, located in another region of Jutiapa, demonstrated promising results [35], suggesting potential interruption of disease transmission with high community acceptance. However, similar approaches in Comapa have not yielded the same success observed in La Brea. Another factor that could affect the persistence of *T. dimidiata* in this region is insecticide resistance, as observed in other regions and other triatomine species [36,37]. Since in Comapa IRS is applied by MoH personnel very focalized and infrequentl, this may not be a main driver of re-infestation. This could be due to a bounce back of sylvatic populations dispersing to the domicile. This refractoriness to both traditional and ecological control strategies highlights the need for sustained vector surveillance and novel control efforts in this region, as complete interruption of vector-borne transmission may be unfeasible, similar to other areas in the Americas [38]. We also observed that the mean density of households with triatomines remained below one in all but one community, comparable to values reported in the 2011 survey[13]. Interestingly, the values of 2011 were observed when communities had been recently intervened with focalized spraying, something also seen by Hashimoto *et al* [39]. It has been previously shown that both *Rhodnius prolixus* [40] and *T. infestans* [41] have much higher household density levels than *T. dimidiata,* which may serve as a more precise indicator of Chagas disease transmission risk. Our findings suggests that even in communities with persistent high infestation and colonization, household vector density may remain low enough to reduce the risk of Chagas disease transmission. This is supported by the observed decline in seroprevalence among school aged children over the years [13], despite not reaching the 8% infestation threshold thought needed for disease transmission interruption [42].

*Trypanosoma cruzi* infections were observed across all life stages of triatomines, as early as first instars. Our results show comparable infection rate as the one observed in the 2011 baseline when 31% of triatomines were positive[9]. This infection rate has also been observed in other rural communities from the southern region of Mexico, were limited control has been applied since it is not considered an endemic disease for that region[43]. Importantly, the infection rate remains below what is generally observed in South American countries were infection prevalence in triatomines can reach up to 68% [44,45].

In our baseline study over a decade earlier in these communities, seroprevalence in 80 adult dogs was 37% [9], compared to 16% in the present study, although different diagnostic approaches adds complexity to assessment of change over time. We observed that one-third of all dogs infected were less than 12 months of age. Given the limited known travel of these dogs, these data suggest active transmission in the household, more importantly these could serve as household reservoirs which ultimately pose a risk for continued human transmission.

Our environmental DNA testing showed that a simple swab from a house wall or floor may be processed to show evidence of the *T. cruzi* parasite, likely by picking up remnants of parasite from areas where infected triatomines have defecated. Given swabbing a home requires no specialized training, such an approach could augment traditional methods of manual triaomine searching of homes. A mixture of traditional and more novel approaches should be considered to improve surveillance methods[18,46], ultimately providing us with in-depth understanding of the ecology of triatomine species.

In our previous study from 2011 in these rural Guatemalan communities, we showed by PCR that the bloodmeal frequency for triatomines was highest for *G. gallus* (64%), followed by humans (50%), dogs (17%), *Rattus rattus* (24%) and *Mus musculus* (21%) [9]. In the current study, we applied a more sensitive approach of bloodmeal metabarcoding, and revealed a similar host community composition over a decade later. *T. dimidiata* is an opportunistic feeder and bloodmeals have been shown in different studies to include all classes of vertebrates [47–49]. The *T. dimidiata* examined in this study showed eight vertebrate hosts with a majority of meals including *Gallus gallus* and *Canis lupus familiaris*, suggests that they robustly support *T. dimidiata* in these environments. A total of five human meals were observed, three were solely human, and two with the presence of other hosts as well.

This predominance of chicken and dog feeding in a population of bugs that also feeds on humans underscores the need for targeted interventions that may include both *Gallus gallus* and *Canis lupus familiaris* that support the life cycle of *T. dimidiata* in the household. For example, chickens have been targeted with systemic indecticides to reduce mosquito populations that can transmit West Nile virus [50]. Dog infection rates and frequency of human bloodmeals in triatomines could in the future be used as outcome variables to analyze the impact of interventions in the short term.

The detection of acute Chagas disease continues to be one of the most daunting tasks for vector-borne disease surveillance programs. The difficulty of finding these cases relies on the direct ability of health care personnel in endemic areas to be aware of the disease, its symptoms and physical manifestations, making this an underdiagnosed type of case [51]. Nonetheless, help from different levels of health providers can greatly increase the chances of finding such a case if disease awareness has been promoted at these levels. In this specific scenario health promoters of this region were a key component for the prompt and correct identification of multiple acute Chagas cases in remote villages of Guatemala. Since 2019, Comapa has established a dedicated Chagas disease clinic to provide medical assistance and follow-up for affected patients. Acute cases are the tip of the iceberg for vector-borne disease transmission where high infestation with triatomines remains, mainly because most cases are asymptomatic and the few that present symptoms may resolve without seeking health care. Improving awareness, detection and laboratory diagnostics of acute cases is critical to provide immediate treatment to patients, which can result in up to 80% of treatment efficacy [51]. However, it is important to recognize that seropositivity can persist long after successful parasite clearance, complicating treatment outcome evaluations. Studies have shown that in adults treated with nifurtimox, less than 60% achieved seronegative conversion even after five years [52], highlighting that seroreversion is a slow process rather than an indicator of treatment failure. In pediatric cases, this pattern is also observed. A study on *T. cruzi*-infected children treated with nifurtimox found that while PCR testing demonstrated parasitological clearance within three years post-treatment, conventional serology remained positive for most individuals beyond this period [53]. This shows the clear need to sustain vector control programs that rely on the use of insecticide and non-insecticide-based approaches to reduce vector-human contact [10]. Additionally, given the prolonged time required for seroreversion, monitoring vector populations and reinfection potential in endemic regions should remain a priority to ensure that achievements made through treatment are not undermined by persistent transmission cycles.

We acknowledge that community sample size in the current study hinders extrapolation to broader geographic regions as does not allow a robust test of the long-term impact of the historic eco-bio-social intervention from 2011. However, this evaluation provides clear and robust information of the ecological patterns of *T. dimidiata* infestation, infection and bloodmeal source in these communities. Additionally, contemporary molecular approaches, including the household eDNA sampling and bloodmeal metabarcoding, provide deeper insight to patterns of infestation and host sources for triatomines. Interestingly, this bloodmeal analytic procedure allowed us to capture preseumed triatomine feeding event on an *Archimandrita* sp.; specimens of this large bodied cockroach were observed in several households across the study sites (Supplementary Results), which showcases the opportunistic feeding nature of *T. dimidiata*.

## Conclusion

The persistence of *Triatoma dimidiata* infestation and *Trypanosoma cruzi* transmission in Comapa, Jutiapa, underscores the complexity of Chagas disease control in endemic regions. Despite multiple intervention strategies over the past decade, infestation rates remain high, and *T. cruzi* infection continues to be detected in both vectors and domestic reservoirs, such as dogs. The detection of human bloodmeals in triatomines highlights an ongoing risk of transmission, reinforcing the need for innovative and sustainable vector control measures. Additionally, the follow-up of acute Chagas disease cases emphasizes the importance of long-term patient monitoring and strengthened diagnostic and treatment strategies. Our findings suggest that while conventional vector control methods have had some impact, more comprehensive and adaptive approaches—including ecological, community-based, and integrated vector management strategies—are required to mitigate the risk of Chagas disease transmission in this region. Continued surveillance and collaboration with local stakeholders will be essential to developing effective and sustainable solutions for Chagas disease prevention and control.

## Supporting information

Supplemental Results

## Data Availability

Data availability
Data set is available in ZENODO: 10.5281/zenodo.14989960, Dryad: https://doi.org/10.5061/dryad.pzgmsbczg and R code can be found at: https://github.com/jgjuarez/Triatoma_2022

https://zenodo.org/records/14989960

https://doi.org/10.5061/dryad.pzgmsbczg

https://github.com/jgjuarez/Triatoma_2022

## Acknowledgements

This study was supported by Fulbright US Scholars Program to SAH, NIH R21AI166446–01 to GLH, and Texas A&M AgriLife Research. We appreciate the support and hospitality provided by members of the community of Comapa during household visits. Nicole Scavo and David Aguilar assisted with household visits and Isabel McAllister assisted with laboratory diagnostics.

## Data availability

Data set is available in ZENODO: https://zenodo.org/records/14989960, Dryad: https://doi.org/10.5061/dryad.pzgmsbczg and R code can be found at: https://github.com/jgjuarez/Triatoma_2022

## Author contribution

J.G.J., E.B., L.M., C.C.R., P.P., N.P., G.L.H and S.A.H. designed and guided the experiments. J.G.J., A.M.M.V., M.G.P., L.A., H.E, R.T., G.L.H. and S.A.H. carried out the data collection and field activities. J.G.J., S.B., L.D.A., E.B., L.A., R.T. analyzed the data. J.G.J.generated the figures. J.G.J., G.L.H. and S.A.H. wrote the manuscript. A.M.M.V., M.G.P., P.P., N.P., S.B., L.D.A., E.B., L.A., H.E., R.T., L.M., C.C.R. reviewed and edited the manuscript. All authors approved the final manuscript.

