## Supplemental Results for "*Triatoma dimidiata,* domestic animals and acute Chagas disease: A 10 year follow-up after an eco-bio-social intervention"

Files included:

- SResults
- SFigure 1

### SResults

#### El Anonito: *T. dimidiata* collection and infection with *T. cruzi* TcI strain

The findings from a home in El Antonio are included as supplemental as this home was not part of the follow-up study. A total of 29 *T. dimidiata* specimens were collected in this household, with 28 nymphs and 1 adult. Specimens were collected in the peridomicile, in a storage room for firewood. All nymphs from 1$^{st}$ (7), 2$^{nd}$ (9) and 3$^{rd}$ (3) instars were processed as described in the main manuscript. We collected one fourth instar nymph, 2 fifth instar nymphs and 1 adult male which were allowed to develop to use for colony establishment at UVG medical entomology laboratory. We detected 15 *T. cruzi* positive triatomines, of which 13 were determined as TcI strain. Infection was detected as early as 1$^{st}$ instar, with 2$^{nd}$, 3$^{rd}$ and 4$^{th}$ also being *T. cruzi* positive.

#### El Anonito: Bloodmeal analysis

We detected a positive bloodmeal from 14 of the 25 nymphs stored in the field. The most frequent meal was *Rattus rattus* (12 one-host bloodmeals and 2 two-host bloodmeals) followed by *Gallus gallus* (1 one-host and 1 two-host bloodmeals). We detected a 2$^{nd}$ instar with a positive human bloodmeal with a dual feeding with *Rattus rattus*, this specimen was also positive for *T. cruzi*.

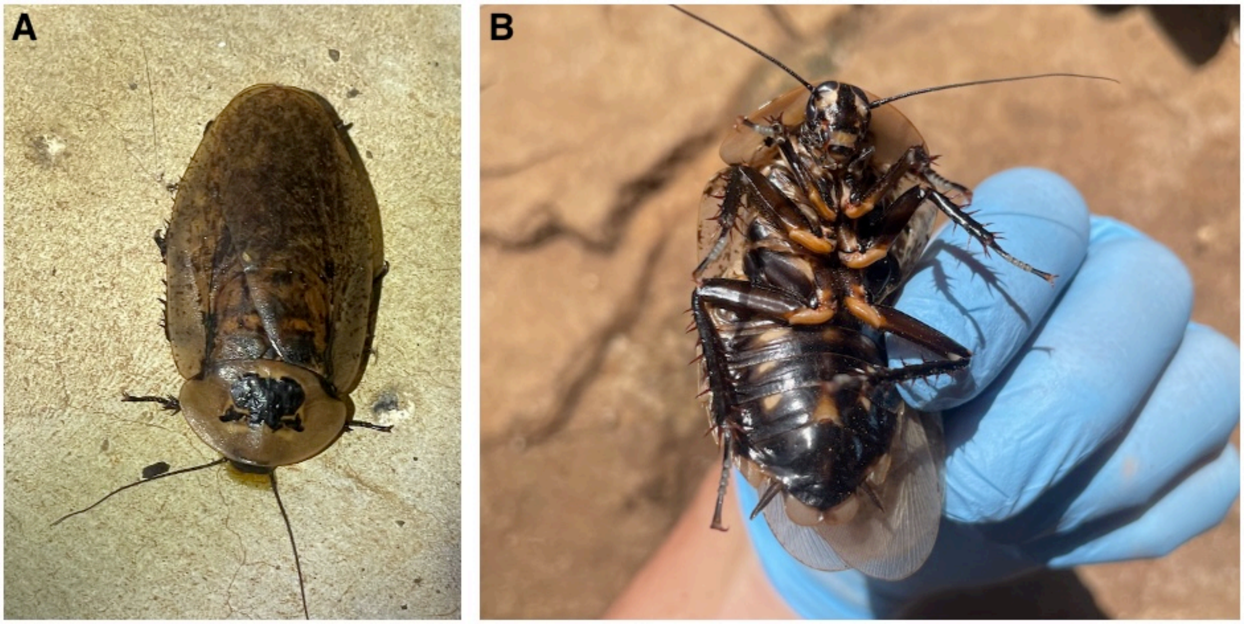


**SFigure 1.** *Archimandrita* sp. cockroach found in a household of Comapa, Juatiapa were *T. dimidiata* was collected. A) Dorsal view. B) Ventral view. *Archimandrita* sp. was detected in the bloodmeal analysis procedure of triatomines suggesting triatomine feeding on hemplymph of this species.
